# Diagnostic Accuracy of CSF Tap-Test Parameters in Predicting Shunt Responsiveness in Normal Pressure Hydrocephalus: A Cohort Study

**DOI:** 10.1101/2025.08.30.25334664

**Authors:** Sagar Poudel, Deepa Dash, Alfonso Fasano, Ajay Garg, Ashish Dutt Upadhyay, Naveet Wig, Roopa Rajan, Animesh Das, MR Divya, Manjari Tripathi, Achal K. Srivastava, SS Kale, P Sarat Chandra, Ashish Suri, Pramod K Pal, Hrishikesh Kumar, Sakoon Saggu, Deepti Vibha, Rajesh Kumar Singh, Jasmine Parihar, Ranveer Singh Jadon, Ved Prakash Meena, Bindu Prakash, Arunmozhimaran Elavarasi

## Abstract

**Background:** The cerebrospinal fluid tap test (CSF-TT) is widely used as an ancillary test to select patients with idiopathic Normal Pressure Hydrocephalus (iNPH) for surgical management; however, its diagnostic utility remains unclear. We evaluate the diagnostic utility of CSF-TT in predicting outcomes following shunt surgery.

**Methods:** Patients with possible or probable iNPH underwent assessments of gait, urinary, and cognitive function using Boon’s gait scale, iNPH grading scale, TUG score, MoCA, and modified Rankin Scale (mRS) at baseline and 24 hours after CSF-TT. They were offered VP shunt surgery based on clinico-radiological profile, regardless of CSF-TT outcome. Post-operative outcomes were evaluated at 24 weeks, and those who showed a ≥1-point improvement in the mRS were classified shunt responders. The diagnostic performance of various scales was assessed using receiver operating characteristic curves.

**Results:** Out of 24 patients who underwent shunting 15 (62.5%) were classified as shunt responders. A one-point reduction in the modified Rankin scale at 24 hours post-CSF-TT had a sensitivity of 53.3% (95% CI: 26.6-78.7) and specificity of 66.7% (95% CI: 29.9-92.5) in predicting shunt responsiveness. Changes in the iNPH score, TUG test score, percentage change in TUG score, Boon’s gait score, and percentage change in Boon’s gait score were not valid predictors since the confidence intervals of the AUROCs crossed 0.5.

**Conclusion:** CSF-TT parameters show limited accuracy in predicting shunt responsiveness in patients with iNPH. Our results suggest that about one-third of positive tap tests were false positives, and the test’s sensitivity was only slightly better than random chance in predicting post-operative shunt outcomes.

## Introduction

Normal Pressure Hydrocephalus (NPH) is characterized by a triad of gait disorder, cognitive impairment, and urinary incontinence. It commonly affects older adults and is equally common in all sexes.^1^ The underlying pathophysiology of idiopathic NPH (iNPH) is not clearly understood; however, it is considered to be due to alterations in CSF dynamics caused by a multifactorial process.^3^ The patients can be categorized into possible NPH, probable NPH, and definite NPH as per Japanese guidelines.^3^ The patients with possible NPH usually undergo CSF tap test (CSF-TT) with monitoring of gait parameters and cognition scores, and improvement in symptoms is often considered a prompt for CSF shunting, i.e., ventriculoperitoneal or lumbo-peritoneal shunt.

There is wide variability in clinical practice and a lack of optimal cutoffs and timing for assessing the CSF-TT to predict improvement after shunt surgery.^2^ Japanese guidelines recommend evaluation after 24 hours following the CSF-TT, with multiple evaluations to be performed over a week to assess expected improvement in cognition and urinary dysfunction.^3^ In the NHS Trust in London, UK, a negative CSF TT is followed by a 48-hour lumbar drainage or a lumbar infusion test, and surgical treatment is offered if either of these tests is positive^3^. In the United States, a review of the National Inpatient Database revealed variations in the protocols for lumbar puncture, external ventricular drain, and CSF diversion methods among different regions.^4^ In Canadian practice, a timed gait assessment is performed after a large volume CSF-TT, and improved gait is considered a positive predictor of response to a ventriculoperitoneal shunt^5^. There is no standard protocol for the performance and assessment of the CSF-TT.^6^ Studies report conflicting results regarding the sensitivity and specificity of CSF-TT.^7–10^ Furthermore, a nationwide survey conducted among Indian neurologists revealed significant heterogeneity in the evaluation and management practices, particularly concerning the utilization of cerebrospinal fluid (CSF) tap test parameters for diagnosis and management of NPH.^11^This study aims to evaluate the diagnostic accuracy of the CSF-TT in predicting outcomes after post-shunt surgery.

## Materials and methods

We conducted an ambispective cohort study of patients presenting with clinical features suggestive of NPH at a tertiary care center in northern India from 2019 to 2024. We recruited all consecutive patients with possible iNPH from 2023 to 2024 prospectively. We retrospectively collected data on patients with iNPH admitted to the neurology ward from 2019 to 2023 by reviewing their charts from the medical records section. The study was approved by the Institutional Review Board. The study was desiged and reported using the STARD reporting guidelines.

Patients who had possible NPH according to Relkin guidelines^12^, with at least one of the specific triad of symptoms of NPH - gait disturbance, urinary disturbances, or cognitive impairment, attending the neurology outpatient department (OPD) were included. We excluded patients with secondary NPH following other causes such as meningitis or subarachnoid hemorrhage. Clinical details, baseline investigations, associated comorbidities, and other symptoms were obtained. Neuroimaging findings were reviewed, and data were extracted into standardized case record forms by a board-certified neuro-radiologist (AG), who was unaware of the results of the CSF-TT. Based on a review of the history, examination, and imaging findings, we excluded patients whose symptoms could be explained by other neurological diseases (Figure 1). Patients with clinical and imaging findings suggestive of NPH were considered for a large-volume lumbar puncture, i.e., CSF-TT, as per the Institute protocol (Box 1). Patients who had probable iNPH, as per the guidelines, were referred for CSF diversion. The Neurosurgery team inserted a programmable Ventriculo Peritoneal shunt (VP Shunt).

**Figure 1:**
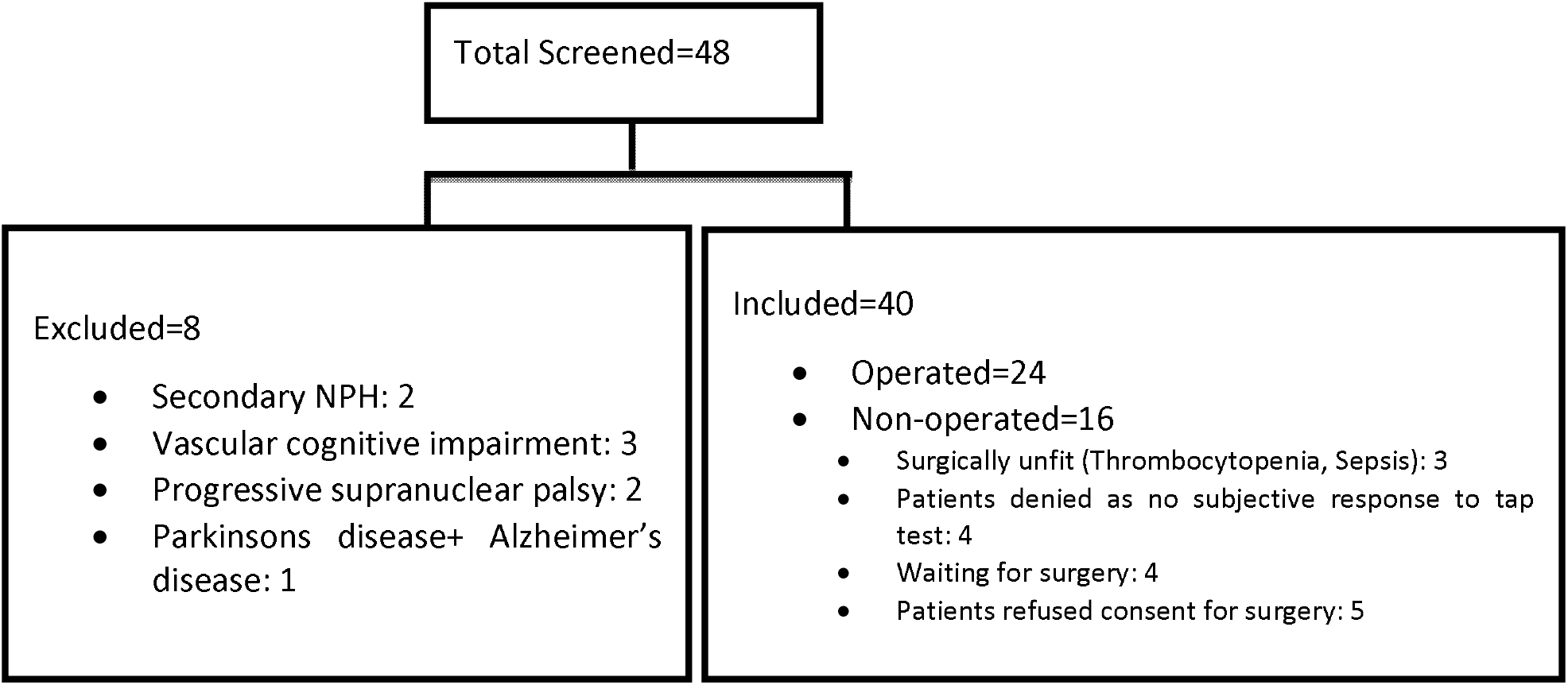
Patients screened and included in the study

### Box 1

**Institute Protocol for the large volume CSF tap test (CSF-TT)the large volume CSF tap test (CSF-TT)**

- Baseline assessment: Less than 24 hours before the CSF-TT
  ○ Gait assessment: Boon’s Gait scale, TUG test, iNPH gait scale, modified Rankin Scale, Videography
  ○ Cognitive assessment: iNPH cognitive scale
  ○ Urinary function: iNPH urinary scale
- Lumbar puncture in the left lateral position
- CSF opening pressure measured with a manometer, after extending the legs
- 30-50 mL of CSF drained, sent for microscopy, biochemistry, and other investigations as dictated by the clinical profile
- Post lumbar puncture assessment of parameters – at 24 hours
- Assessed using scales done at baseline, repeated at 24 hours
- 3, 6-hour, and 12-hour values are recorded as well
- Cognitive assessment was performed using the MoCA only in the prospectively recruited patients.

### Post-operative outcome

A follow-up assessment was performed post-operatively, including a general assessment of gait, cognition, and improvement in urinary symptoms, as well as documentation of the iNPH grading scale score and the mRS at 24 weeks. Telephonic interviews were conducted to assess the patient or caregiver recall based mRS at 24 weeks in retrospectively recruited patients. In this study, we defined shunt responsiveness as a reduction of at least 1 point on the modified Rankin scale at 24 weeks after the Ventriculoperitoneal shunt surgery. In the prospectively recruited patients, we also used the post-operative iNPH grading scale, and a one-point reduction in the iNPH scale was considered indicative of shunt responsiveness. The assessments were conducted either in-clinic during follow-up visits or telephonically, as per the patient’s preference.

### Statistical analysis

Data were collected and managed using REDCap (Research Electronic Data Capture), a secure, web-based application designed to support data capture for research studies. The REDCap platform used in this study was hosted by the All India Institute of Medical Sciences (AIIMS), New Delhi, India. Data were exported and cleaned using Microsoft Excel (Microsoft Corp., Redmond, WA, USA). Descriptive statistics, including frequencies and percentages, were used to describe categorical variables. Mean and standard deviations were used for continuous variables with a normal distribution, while medians and ranges were used for non-parametric data. For comparisons between groups, an independent samples t-test was used for normally distributed data, and the Mann–Whitney U test was used for non-normally distributed data. Dichotomous variables were compared using the Chi-squared test or Fisher’s exact test. ROC curve analysis was used to evaluate the ability of the CSF-TT to predict shunt responsiveness. The area under the curve (AUC) was computed, and the optimal cutoff was determined using the Youden index. Sensitivity and specificity were calculated at the chosen threshold. All statistical analyses were performed using Stata version 17 (StataCorp, College Station, TX, USA). A p-value of <0.05 was considered statistically significant.

## Results

We screened 48 patients: 20 in the retrospective arm (2019-2022) and 28 in the prospective arm. Eight patients were excluded (Figure 1), and 40 patients underwent a CSF-TT. Out of 40 patients considered for a CSF-TT, only 24 underwent VP shunt surgery. Among the 24 patients, 21 had all three symptoms of the triad, and 3 had at least two symptoms of the triad.

The shunt responders were significantly older than the non-responders. The duration of symptoms before diagnosis was similar between the two groups. However, the duration of cognitive impairment was longer in non-responders (24 months vs. 12 months in responders), but this difference was not statistically significant. The MoCA scores were significantly lower in the non-responders (Table 1). There were no differences between the groups in the iNPH score or modified Rankin scale at baseline. The baseline characteristics of the shunt responders and non-responders have been summarized and compared in Table 1.

**Table 1:** Baseline characteristics

| Variable | Shunt Responders<br>(n=15) | Non-Responders<br>(n=9) | p value |
| --- | --- | --- | --- |
| Age (years), mean± SD | 67.2 ± 4.3 | 58.9 ± 13.5 | 0.04 |
| Male Sex, n(%) | 13 (86.7%) | 7 (77.8%) | 0.61 |
| Gait Disturbance, n(%) | 15 (100%) | 9 (100%) | - |
| Urinary Disturbance, n(%) | 14 (93.3%) | 9 (100%) | >0.99 |
| Cognitive Impairment, n(%) | 13 (86.6%) | 9 (100%) | 0.51 |
| Parkinsonism(n=11), n(%) | 6(40%) | 5(55.5%) | 0.68 |
| Lower body Parkinsonism (n=4), n(%) | 2(13.3%) | 2(25%) | 0.68 |
| Falls (n=11), n (%) | 6(40%) | 5(55.5%) | 0.68 |
| Duration of Symptoms (months), median (IQR) | 26 (12–84) | 24 (11–60) | 0.59 |
| Gait dysfunction duration (months), median (IQR) | 24(12-84) | 24 (9-60) | 0.51 |
| Cognitive impairment duration (months), median (IQR) | 12(5-36) | 24(6-48) | 0.07 |
| Urinary symptom duration(months), median (IQR) | 18.5(5-60) | 12(1-78) | 0.49 |
| MOCA score (n=7), mean± SD | 20.8± 2.2 | 9±2.8 | <0.01 |
| TUG test (n=10), mean±SD | 27.5±10.2 | 43±1.4 | 0.08 |
| iNPH Composite Score, median (IQR) | 4 (3-7) | 5 (3-7) | 0.88 |
| Duration to shunt after CSF-TT (days), median (IQR) | 22 (14-79) | 15 (11-50) | 0.47 |
| modified Rankin score, mean (SD) | 3.3± 0.8 | 2.9± 0.9 | 0.30 |
| mRS 1 n(%) | 0 | 0 | 0.22 |
| mRS 2 n(%) | 2(13.3%) | 4 (44.4%) |  |
| mRS 3 n(%) | 8 (53.3%) | 2 (22.2%) |  |
| mRS 4 n(%) | 4 (26.7%) | 3 (33.3%) |  |
| mRS 5 n(%) | 1 (6.7%) | 0 |  |

We compared the change in the scales used for assessment in Table 2. There were no significant differences between groups in terms of change from baseline to post CSF-TT as assessed by the modified Rankin scale, iNPH composite score, Boon’s step score, or the Boon’s total gait score. However, there was a significant reduction in the Boon’s time score after CSF-TT in the shunt responders compared to non-responders. Out of the eight parameters of gait as assessed by Boon’s gait score, significantly more patients in the shunt-responder group had improved foot clearance after CSF-TT as compared to non-responders. Interestingly, out of three patients who were unable to walk without assistance at baseline, two showed improvement in their gait and were able to walk without support after the tap test. However, they did not respond to the shunt surgery. We found no significant difference in the time to VP shunt surgery after the tap test between responders and non-responders (median 22 days, IQR 14–79 vs. 15 days, IQR 11–50; p = 0.47). Surprisingly, one patient who was unable to walk at all at baseline and showed no improvement after the tap test was able to walk unassisted post-surgery.

**Table 2:** Change in Assessment Scales 24 hours after the CSF-TT

| Parameter | Shunt responder (n=15) | Shunt non-responder (n=9) | P value |
| --- | --- | --- | --- |
| Change in Boon's Walk Score, median(IQR)* | 4 (0-6) | 2 (2-4) | 0.85 |
| **Change in the components of Walk Score<sup>#</sup>** |  |  |  |
| Improvement in tandem walking, n/N(%) | 2/12 (16.7%) | 1/6 (16.7%) | >0.99 |
| Improvement in turning, n/N(%) | 6/12 (50%) | 3/6 (50%) | >0.99 |
| Improved Trunk Balance, n/N(%) | 1/12 (8.3%) | 1/6 (16.7%) | >0.99 |
| Improvement in Wide-Based Stride, n/N(%) | 5/12 (41.7%) | 0 | 0.11 |
| Improvement in Small Steps, n/N(%) | 7/12 (58.3%) | 1/6 (16.7%) | 0.15 |
| Improved Foot Clearance, n/N(%) | 7/12 (58.3%) | 0 | 0.04 |
| Improved Start Hesitation, n/N(%) | 4/12 (33.3%) | 2/6 (33.3%) | >0.99 |
| Resolved Tendency to Fall, n/N(%) | 1/12 (8.3%) | 1/6 (16.7%) | >0.99 |
| Able to walk unassisted from pre-tap assisted walking/inability to walk, n/N(%) | 0 | 2/6 (66.7%) | 0.4 |
| Steps score change, median (IQR)* | 2 (1-3) | 1 (0-2) | 0.09 |
| Time score change, median (IQR)* | 2 (0-2) | 0 (0-0) | 0.02 |
| Gait Scale Total Score change, median (IQR)* | 10 (3-11) | 4 (2-5) | 0.51 |
| MoCA change, median(IQR)* | 1 (1-3) | 5 (0-0) | 0.10 |
| TUG change, median(IQR)* | 5 (2-10.5) | 15 (10-20) | 0.15 |
| iNPH Composite Score change, median (IQR)* | 1(0-1) | 1(0-1) | >0.99 |
| Modified Rankin Score change, median (IQR)* | 1(0-1) | 0 (0-1) | >0.99 |
| No change in mRS n(%) | 7 (46.7%) | 6 (66.7%) | 0.46 |
| mRS improvement by 1 point, n(%) | 7 (46.7%) | 2 (22.2%) |  |
| mRS improvement by 2 points, n(%) | 1 (6.7%) | 1 (11.1%) |  |
| * Change in continuous scales: Score before CSF-TT – Score assessed 24 hours after the CSF-TT |  |  |  |
| * Change in dichotomous outcomes: Those that improved from not being able to do the gait task before CSF-TT to being able to do the gait task after |  |  |  |

Among the 24 patients who underwent shunt surgery, 15 were responders. In this group of shunt responders, seven were tap test non-responders, while eight patients responded to the CSF-TT. However, three patients who were CSF-TT responders died before 24 weeks’ follow-up, and six patients who were tap test non-responders did not respond to the shunt as well. (Table 2) Our findings show that only 62.5% responded to VP shunt surgery at 24-week follow-up. Thus, the prevalence of definite NPH in a highly selected cohort of patients with clinico-radiologically probable iNPH (irrespective of the CSF-TT results) was 62.5%.

To evaluate the ability of CSF-TT parameters to predict shunt responsiveness, we assessed the diagnostic performance of various assessment scales to determine shunt responsiveness 24 weeks after shunt placement (Table 3). At least one point reduction in mRS following CSF tap showed a sensitivity of 53.3% (95% CI: 26.6%-78.7%) and a specificity of 66.7% (95% CI: 29.9%-92.5%). A greater than 10-point improvement in Boon’s gait score following the CSF-TT provided the best discriminatory value for predicting shunt responsiveness, with a sensitivity of 53.3% and specificity of 77.8%, yielding the highest Youden Index. However, a 37% increase in the Boons score resulted in increased specificity at 88.9% with the highest AUROC of 0.66 (95% CI: 0.42-0.89) (Figure 2). ROC analysis of TUG score changes after CSF tap yielded an AUROC of 0.16 (95% CI: 0.00–0.50), indicating very poor discriminatory ability for predicting shunt responsiveness in our cohort. None of them were statistically significant in predicting shunt responsiveness. We also evaluated the diagnostic performance of these parameters in predicting shunt responsiveness, defined as a one-point improvement in the iNPH score 24 weeks after surgery. The reduction in iNPH score by at least 1 point after CSF tap had a sensitivity of 64.3% (95% CI: 35.1%-87.2%) and a specificity of 66.7% (95% CI: 22.3%-95.7%). The results were no different from those obtained using the modified Rankin scale to define shunt responsiveness. The results are summarized in Table 3.

**Table 3:**
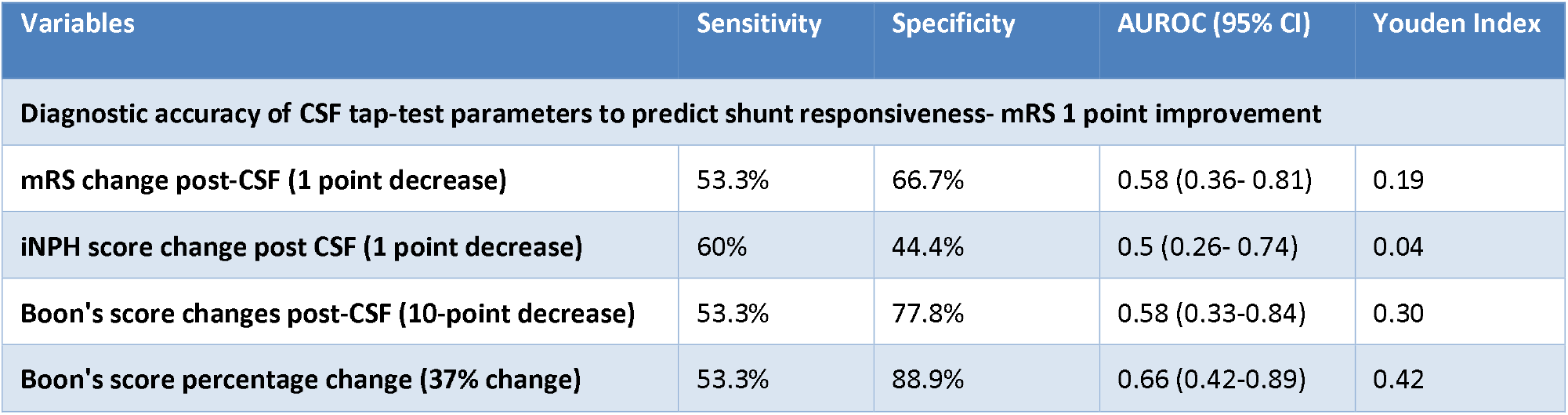
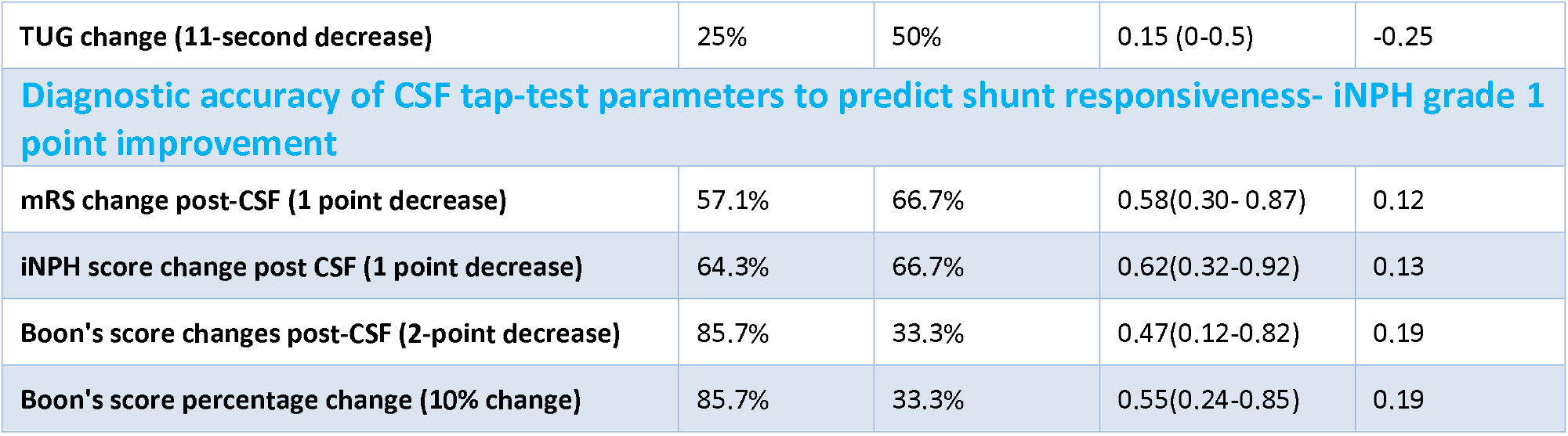
Diagnostic accuracy of CSF tap-test parameters to predict shunt responsiveness

**Figure 2:**
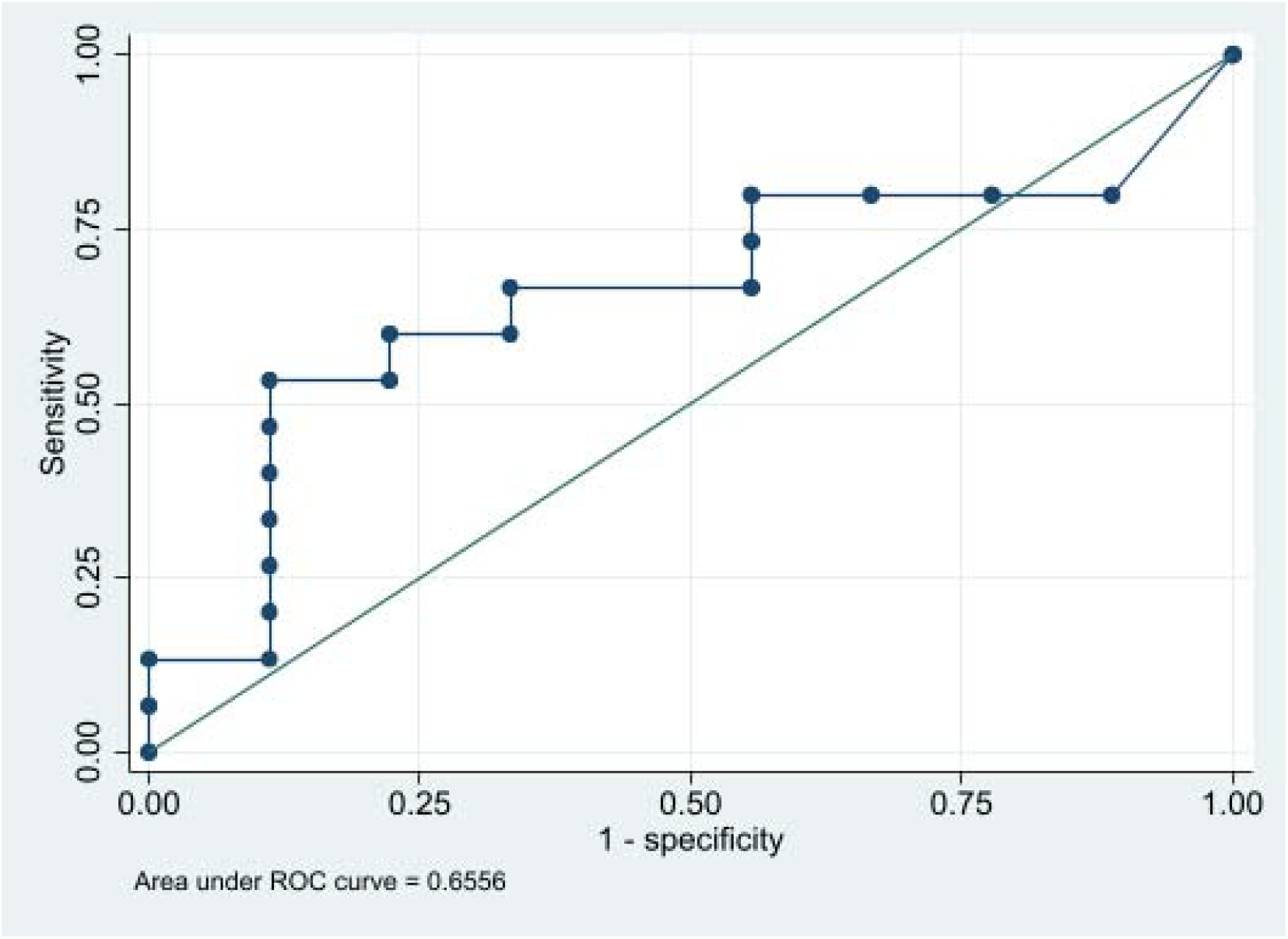
ROC curve for predicting reduction in mRS post-Ventriculoperitoneal shunt with percentage Boon’s score change post-CSF-TT (95% CI: 0.42-0.89)

## Discussion

In this cohort study, we aimed to investigate the diagnostic utility of CSF-TT parameters in predicting shunt responsiveness in patients with iNPH. We used the mRS as the primary outcome measure in our study. Most studies on idiopathic NPH have used the iNPH scale to assess outcomes. We found that the modified Rankin scale had a higher Youden index as compared to the iNPH scale (Table 3)

Unfortunately, patients often experience symptoms for a considerable duration, typically 24 to 26 months, before getting a diagnosis. Despite this delay in diagnosis, we found that almost two-thirds showed significant improvement at 24 weeks post-VP shunt. This is slightly lower compared to the study by Hashimoto et al., where an 80% improvement was observed using mRS.^13^ It is possible that if patients are identified and treated early, the outcomes may differ, and thus the diagnostic accuracy may also be different. In our study, we found that shunt responders were significantly older compared to non-responders, which was surprising, as older age is usually associated with poorer prognosis.^14^ Although results from the SINPHONI study did not reveal an age difference between the groups.^9^ Also, a significant difference was observed in the baseline MoCA scores between shunt responders and non-responders, with higher MOCA scores in the shunt responders signifying a lesser degree of cognitive impairment. This is another important finding that might help prognosticate patients before surgery. However, studies have shown that cognitive impairment does not differ between the groups.^9,15^

CSF-TT is considered an important ancillary test for predicting shunt response in patients with NPH. The TUG test is a commonly used assessment tool to evaluate gait. The diagnostic performance of changes in TUG scores post-CSF-TT in predicting shunt responsiveness was very poor (Sensitivity 25% and Specificity 50%). This finding aligns with a study where a >10% improvement in TUG test score was associated with a low sensitivity of 34.2% and a specificity of 73.6%.^9^ This highlights the limited utility of the TUG test in the diagnosis and management of NPH.

While Boon’s gait scale is widely used for qualitative gait assessment, there is limited literature supporting its utility specifically for the tap test in predicting shunt responsiveness. A detailed analysis of gait parameters among patients who walked independently revealed that turning disturbance, small steps, reduced foot clearance, and wide-based strides were the most commonly improved gait parameters among shunt responders. Improvement in foot clearance was significantly higher among shunt responders. This contrasts with the study where the tendency toward falling and tandem walking disturbance was the most improved gait parameter after CSF-TT.^16^ Similarly, in a study by Ravdin et al, improvement in turning, trunk balance, tandem walking, start hesitation, and tendency towards falling showed maximum improvement after CSF-TT^17^. This highlights the variability and subjectivity of gait parameters in assessing responsiveness to the tap test. This also questions the validity of each subscale of the Boon’s gait scale and the concept of adding each subcomponent to create the total walk score. The Boon’s gait score ranges from 0 to 40, with 10 points allocated to the step score, 10 points to the time score, and 20 points to the walk score, which comprises eight subcomponents. If the patient is unable to walk independently, the walk score is 18 if they can walk with assistance and 20 if the patient is unable to walk at all. This score suffers from a lack of discrimination between a patient who cannot walk at all and someone with the highest possible score of 10 on the time and step score, both of whom will be scored 20 points each. This thereby showcases the poor discriminatory capacity even at baseline.

Various studies have used changes in gait velocity, step length, stride length, TUG test score, and iNPHGS to determine response to the tap test. Some studies have used absolute reductions in values, while others have used percentage changes. ^13,18^ Our results suggest that CSF-TT parameters, including changes in the iNPH composite score and Boon’s gait scale, as well as the TUG score, have limited predictive value for shunt responsiveness. We also found significant discordance in response to CSF TT and shunt responders, as assessed by the modified Rankin scale, and vice versa (Table 2). Compared to the SINPHONI studies^13^, which reported higher sensitivity (71–83%) and specificity (65–85%) using broader clinical and imaging criteria, our study yielded lower values. This discrepancy may reflect differences in methodology, tap test volume, timing of assessments (fixed at 24 hours in our study), and outcome definitions (functional outcome using mRS versus iNPHGS or TUG test)^8,19^. The sensitivity and specificity depend on the cutoffs assumed for test positivity and thresholds for shunt responsiveness. In the SINPHONI study, the specificity was 80% and the sensitivity was 51.3% using gait change in iNPHGS^9^. In our recent meta-analysis^20^ of eight studies, the pooled sensitivity of the CSF-TT is 66.5% (95% CI: 49.6-81.6), and the pooled specificity is 52% (95%CI: 35.7-68.1). This emphasizes on the fact that the clinical utility of CSF-TT is analogous to tossing a coin.

This study has several limitations. First, the sample size was relatively small, with only 24 patients undergoing shunt surgery, which may limit the statistical power and generalizability of the findings. Out of 40 patients who fulfilled the clinical and radiologic criteria for probable iNPH, only 24 underwent surgery. Though the patients were advised shunt surgery based on the clinico-radiologic profile, some patients refused consent as they did not feel subjective improvement following the CSF-TT, and this could have introduced selection bias. The Ambispective design relied partly on retrospective data, which may introduce recall bias, particularly in caregiver-reported or patient self-reported outcomes during follow-up. Few patients were followed up telephonically, and this could have led to heterogeneity in outcome assessment.

In most cases, the same examiner administered the assessment scales before and after the CSF-TT and was not blinded to the timing of the assessment in relation to the tap test. The TUG test was available only in prospectively collected patients, and thus, the numbers were low and may not have been powered enough to detect meaningful differences. The index test results were available to the outcome assessor. The outcomes were not assessed using objective gait analysis parameters using sensors, which could have impacted the classification of qualitative parts of the scales, especially Boon’s Gait Scale, which, though practical, may lack standardization across observers. Comorbidities, such as orthopedic issues, could have influenced the outcomes; however, to ensure generalizability, functional scores were used to assess outcomes. All patients in our cohort received programmable shunts; however, valve pressure readjustments were not systematically recorded or analyzed. It is possible that such adjustments occurred after the completion of our predefined outcome assessments, which may have influenced the observed functional outcomes. Lastly, the study used a single time point assessment at 24 hours following the CSF-TT, whereas some patients may exhibit delayed improvements, which could have affected the sensitivity of the tap test.

## Conclusion

The overall diagnostic accuracy remains suboptimal, and often no better than tossing a coin to predict shunt responsiveness. Many patients who ultimately benefit from shunting may not show significant improvement on CSF-TT, while others may exhibit transient improvement post-operatively without long-term surgical benefits. One of the significant challenges in the field is the current definition of “definite” iNPH, which is based entirely on post-shunt improvement. This inherently circular definition limits our ability to characterize the disease in its preoperative state and complicates early identification of likely responders. Moreover, the response to shunt surgery is influenced by a range of extraneous variables, including the type and timing of surgical intervention, the type of valve used and valve pressure settings, post-operative complications such as overdrainage or infection, and the patient’s overall systemic health and comorbidities. These factors may obscure or confound the true underlying biology of the disease. Our findings highlight the inherent limitations of relying solely on CSF-TT outcomes to decide intervention or post-surgical response to define definite iNPH. There is a compelling need for a comprehensive, multimodal diagnostic approach that integrates clinical assessment with standardized functional scales, objective gait and cognitive measurements, advanced neuroimaging biomarkers (e.g., callosal angle, white matter integrity, DTI), and possibly molecular or CSF-based biomarkers. Such an approach could enhance diagnostic accuracy, identify likely responders earlier, and tailor treatment strategies to individual patient profiles. Future research must focus on refining diagnostic criteria and identifying surrogate markers that are independent of therapeutic outcomes to advance the field of iNPH care.

## Data Availability

All data produced in the present study are available upon reasonable request to the authors

## Funding sources and conflict of interest

No specific funding was received for this work.

## Conflict of interest statement

The authors declare that there are no relevant conflicts of interest Financial Disclosures: The authors declare that there are no additional disclosures to report.

## Ethical Compliance Statement

The study was reviewed and approved by the Institute Ethics Committee at the All India Institute of Medical Sciences, New Delhi (No. IECPG-222/20.4.23, RT-14/07.06.23). Informed consent to participate in this study was provided by the participants and their legal guardians.

We confirm that we have read the Journal’s position on issues involved in ethical publication and affirm that this work is consistent with those guidelines.

## Legends

Supplementary table 1: Comparison of before and after CSF tap test Boon’s gait parameters, iNPH grading score, and Modified Rankin Scores between shunt responders and non-responders

## Notes

### Competing Interest Statement

The authors have declared no competing interest.

### Funding Statement

This study did not receive any funding

### Author Declarations

Ethical Compliance Statement: The study was reviewed and approved by the Institute Ethics Committee at the All India Institute of Medical Sciences, New Delhi (No. IECPG-222/20.4.23, RT-14/07.06.23). Informed consent to participate in this study was provided by the participants and their legal guardians.

