## Supplemental Table 1 for "Diagnostic Accuracy of CSF Tap-Test Parameters in Predicting Shunt Responsiveness in Normal Pressure Hydrocephalus: A Cohort Study"

**Supplementary table 1: Comparison of before and after CSF tap test Boon’s gait parameters, iNPH grading score, and Modified Rankin Scores between shunt responders and non-responders**

|  | **Baseline (Pre CSF Tap test)** | | **P value** | **Post CSF Tap Test** | | **P value** |
| --- | --- | --- | --- | --- | --- | --- |
|  | **Shunt responder(n=15)** | **Shunt non responder(n=9)** |  | **Shunt responder post tap(n=15)** | **Shunt non-responder post tap(n=9)** |  |
| **Boon’s gait score** | | | | | | |
| **Components of walk score** | | | | | | |
| *Walking Independently, n(%)* | 12(80%) | 6(66.7%) | 0.64 | 12 (80%) | 8 (88.8%) | >0.99 |
| *Tandem Walking, n(%)* | 12(100%) | 5(83.3%) | 0.33 | 10 (83.3%) | 6 (75%) | >0.99 |
| *Turning Disturbed, n(%)* | 8(66.7%) | 5(83.3%) | 0.62 | 2 (16.7%) | 3 (37.5%) | 0.35 |
| *Trunk Balance, n(%)* | 6(50%) | 2(33.3%) | 0.64 | 5 (41.7%) | 2 (25%) | 0.64 |
| *Wide-Based Stride, n(%)* | 7(58.3%) | 3(50%) | >0.99 | 3 (25.0%) | 4 (57.1%) | 0.36 |
| *Small Steps, n(%)* | 10(83.3%) | 5(83.3%) | >0.99 | 4 (36.3%) | 6 (75%) | 0.17 |
| *Reduced Foot Clearance, n(%)* | 12(100%) | 5(83.3%) | 0.33 | 5 (41.7%) | 6 75%) | 0.2 |
| *Start Hesitation, n(%)* | 5(41.7%) | 4(66.7%) | 0.62 | 1 (8.3%) | 3 (37.5%) | 0.26 |
| *Tendency to Fall, n(%)* | 4(33.3%) | 2(33.3%) | >0.99 | 4 (33.3%) | 2 (25%) | >0.99 |
| *Able to Walk assisted, n(%)* | 2(13.3%) | 3(33.3%) | >0.99 | 2(13.3%) | 1(11.1%) | >0.99 |
| *Unable to walk at all, n(%)* | 1(6.7%) | 0 |  | 1(6.7%) | 0 |  |
| **Total Walk Score, mean±SD** | 12.3±4.8 | 12.9±4.6 | 0.76 | 8.3±6.1 | 9.1±5.8 | 0.74 |
| **Steps score, mean±SD** | 7.7±2.3 | 7.8±2.8 | 0.95 | 5.5±2.2 | 6.6±3.2 | 0.35 |
| **Time score mean±SD** | 8.1±2.7 | 7.4±2.6 | 0.54 | 6.4±2.7 | 7.1±2.9 | 0.57 |
| **Boon’s Gait Scale Total Score mean±SD** | 27.1±7.7 | 28.1±9.6 | 0.77 | 19.4±8.6 | 22.8±10.6 | 0.40 |
| **iNPH score** | | | | | | |
| ***iNPH Gait Score, median(IQR)*** | 1(1-2) | 1(1-3) | 0.91 | 1 (1-2) | 1 (1) | 0.94 |
| ***iNPH Cognition Score, median(IQR)*** | 1(1-3) | 1 (1) | 0.33 | 1(1-2) | 1(0-1) | 0.49 |
| ***iNPH Urinary Score, median(IQR)*** | 1(1-3) | 3(1-3) | 0.2 | 1(1-2) | 2(1-3) | 0.23 |
| ***iNPH Composite Score, median(IQR)*** | 4 (3-7) | 5 (3-7) | 0.88 | 3 (2-6) | 5 (2-6) | 0.76 |
| **MOCA score(n=7) mean±SD** | 20.8± 2.2 | 9±2.8 | <0.01 | 22.6±1.1 | 14±2.8 | <0.01 |
| **TUG score(n=10) mean±SD** | 27.5±10.2 | 43±1.4 | 0.08 | 22±6.5 | 28±5.7 | 0.27 |
| **Modified Rankin Score, mean ±SD** | 3.3± 0.8 | 2.9± 0.9 | 0.30 | 2.7± 1.0 | 2.4± 1.1 | 0.62 |
| **mRS 1***, n(%)* | 0 | 0 | 0.22 | 1 (6.7%) | 2 (22.2%) | 0.79 |
| **mRS 2***, n(%)* | 2(13.3%) | 4 (44.4%) |  | 7 (46.7%) | 3 (33.3%) |  |
| **mRS 3***, n(%)* | 8 (53.3%) | 2 (22.2%) |  | 4 (26.7%) | 2 (22.2%) |  |
| **mRS 4***, n(%)* | 4 (26.7%) | 3 (33.3%) |  | 2 (13.3%) | 2 (22.2%) |  |
| **mRS 5***, n(%)* | 1 (6.7%) | 0 |  | 1 (6.7%) | 0 |  |
